# Prediction of Type 2 Diabetes using genomics and proteomics: the HUNT Study (ProtRS for T2D)

**DOI:** 10.64898/2026.09.06.26362374

**Authors:** Fikria Karinanur, Bjørn Olav Åsvold, Guro F. Giskeødegård, Eivind Almaas, Christian Jonasson, Weihong Tang, John-Bjarne Hansen, Kristian Hveem, Ingrid Sørdal Følling, Brooke N. Wolford

## Abstract

Type 2 diabetes (T2D) is a multifactorial metabolic disorder with an increasing global prevalence and significant health burdens. Early identification of individuals at high risk is crucial for timely intervention and prevention of T2D and complications. This study aims to investigate the association between the blood plasma proteome and T2D, identify protein biomarkers associated with incident T2D, and explore the underlying biological pathways involved in the disease development in the Trøndelag Health Study (HUNT).

The HUNT Study is a population-based cohort with four waves of enrolment beginning in 1984. Samples from HUNT3 were analysed with SomaScan including 3,221 using SomaScanv4.0 (HUNT-Soma5k, ∼5,000 proteins) and 2,181 on SomaScanv4.1 (HUNT-Soma7k, ∼7,000 proteins). We performed logistic regression and least absolute shrinkage and selection operator (LASSO) to generate the protein risk scores, and gene set enrichment analysis (GSEA) to identify potential biological pathways for incident T2D.

We identified 313 (308 proteins) and 142 (128 proteins) aptamers that are significantly associated with incident T2D, in HUNT-Soma5k and HUNT-Soma7k, respectively. In HUNT-Soma5k, MXRA8 has strongest association with decreased risk and PLXB2 has strongest association with increased risk; while IGFBP2 has strongest association with decreased risk and INHBC has strongest association with increased risk for incident T2D in HUNT-Soma7k. The most significant pathway from GO Biological Process is organic acid metabolic process in both datasets.

The associated proteins and biological pathways identified represent insights into the underlying molecular mechanism of T2D and may serve as potential biomarkers for risk prediction and prevention for T2D.

**Article Highlights:** Type 2 diabetes (T2D) leads to substantial morbidity, including cardiovascular, renal, and neurological complications. Identifying individuals at elevated risk before clinical onset is essential, as early intervention can prevent progression from pre-diabetes and support timely lifestyle modification. In this study, we evaluated circulating proteins as predictors of future T2D using high-throughput, targeted proteomics. We identified hundreds of proteins associated with incident T2D, including 10 proteins not previously reported in other cohorts. These findings may enhance early risk prediction and inform more personalized prevention strategies.

## Introduction

Type 2 diabetes (T2D) is a global health challenge with a substantial clinical and economic burden^(1)^. Around 90% of diabetes mellitus cases are T2D, and its global prevalence now exceeds 6%, with some regions reporting even higher rates^(1)^. In Norway, the prevalence reached 7.6% in 2021, with a relatively stable incidence during 2009-2021^(2)^. Estimates from population-based studies indicated that over 10% of adults are at high risk for developing T2D, with higher risk observed in women^(3, 4)^.

T2D is associated with reduced life expectancy and increased risk of cardiovascular disease (CVD), chronic kidney disease (CKD), neuropathy, and retinopathy^(5)^. In European cohorts, the prevalence of coronary heart disease, CKD, foot ulcers, and diabetic retinopathy among individuals with T2D ranges from 5% to 29%^(6–9)^. These complications underscore the need for early detection and targeted prevention strategies. Traditional screening tools rely on clinical risk scores and glycaemic markers, which may not capture early biological changes. In Norway, the national diabetes guidelines^(10)^ still cite the prediction tools FINDRISC^(11)^, but in practice, clinicians measure HbA1c if there is clinical suspicion based on risk factors. Similar limitations exist in the UK and across Europe, where population-level screening is not recommended. In contrast, the U.S. Preventive Services Task Force advocates risk-based screening for adults aged 35–70 with elevated BMI, with earlier testing for high-risk ethnic groups^(12)^. Ultimately, there is a need for an objective T2D risk score with high sensitivity and specificity.

Over 600 genetic variants have been associated with T2D, most with modest effect sizes^(13)^. Recent advances in genomics have led to the development of polygenic scores (PGS), which aggregate the effects of these genome-wide genetic variants to estimate disease susceptibility. PGS can improve risk stratification when combined with clinical factors, but their clinical utility remains to be proven^(14)^. Proteomics offers a complementary approach by capturing dynamic biological signals that reflect disease processes. Studies using large cohorts across European, African, African American, and Middle-Eastern ancestries have identified hundreds of circulating proteins associated with prevalent and incident T2D^(15–19)^. These findings suggest that proteomic risk scores (ProtRS) may enhance early detection and provide mechanistic insights beyond genomics alone. This study aims to evaluate the association between circulating proteins and future risk of T2D, leveraging high-throughput, targeted proteomics to identify individuals at risk before clinical onset. By integrating proteomic data with clinical covariates, we create a ProtRS for T2D. Our results seek to improve risk prediction and inform personalized prevention strategies, in the absence of a widely used risk predictor for T2D.

## Research Methods and Design

### The HUNT Study

The Trøndelag Health (HUNT) Study is an ongoing population-based health study in Trøndelag County, Norway^(20–22)^. The study collects health-related data from questionnaires, interviews, and clinical examinations from individuals within this geographical region. The periodic survey design includes four recruitment waves. HUNT1 (1984–1986), HUNT2 (1995–1997), HUNT3 (2006–2008), and HUNT4 (2017–2019) concentrated primarily in the northern region of Trøndelag county, where all adults (age ≥ 20 years) were invited. We used proteomics data from HUNT3. Additional demographic information such as sex, age at time of blood sample, anthropometric measurements, physical activity index, eGFR, and smoking status from questionnaires and clinical measurements was used to account for confounding. The k-nearest neighbour (kNN) method using R package ‘*VIM*’^(23)^, a machine learning method for imputation, was performed to impute missing values and replace outliers in the covariates, such as weight, height, waist circumference, hip circumference, physical activity index, and smoking status for T2D-proteins association analysis.

### Proteomics assay

EDTA-plasma samples from HUNT3 enrolment visit, stored at -80 °C, were used for proteomic profiling. Proteomic analyses for the HUNT-Soma5k cohort were performed in 2017 using the SomaScan v4.0 platform (5,214 aptamers, 4,930 proteins), while analyses for the HUNT-Soma7k case-cohort were conducted in 2022 using the extended SomaScan v4.1 platform (7,289 aptamers, 6,428 proteins). 5,032 aptamers for 4,879 proteins were measured in both assays. The HUNT-Soma5k cohort consists of 3,221 participants with prevalent and incident cardiovascular disease and controls^(24)^. The HUNT-Soma7k consists of 2,181 participants with venous thromboembolism (VTE) or abdominal aortic aneurysm (AAA) and a random sample of HUNT3 as a subcohort for controls^(25)^. 226 participants were in both datasets, however due to the difference of the selection criteria, we performed the analyses separately. Proteins and samples flagged as failing QC were excluded. Protein abundance was log transformed and standardized across samples to mean 0 and standard deviation 1.

### Genotyping and Polygenic Scores

Genotyping and imputation were performed as previously described^(26)^. Briefly, genotyping was done using Illumina HumanCoreExome arrays. Genotype calling followed a Genome Studio quality control protocol. Variants failing mapping, cluster separation, Hardy Weinberg equilibrium, or low call rates were excluded. Imputation was conducted on 69,716 samples with genetic ancestry similar to that of European populations of the Human Genome Diversity Panel using Minimac39 with default settings and a customized Haplotype Reference Consortium^(27)^ release 1.1 (HRC v1.1) for autosomal and chromosome X variants. The customized reference panel incorporated low-coverage whole-genome sequences from the HUNT study and HRC v1.1 data. Imputed variants with Rsq < 0.3 were excluded, resulting in over 24.9 million variants. The polygenic score (PGS) was calculated using the PGS Catalog Calculator^(28, 29)^ for participants with genetic data from HUNT2-4. We used PGS004152^(30)^ which was an ensemble PGS from a T2D GWAS in 26,676 cases and 132,532 controls^(31)^. The dosages for single nucleotide polymorphisms were used for estimating the PGS.

### Type 2 Diabetes definition

We defined type 2 diabetes as the presence of ICD10 code E11 or ICD9 code 250 based on International Classification of Diseases (ICD) codes from electronic health records of the three hospitals in the catchment area. For the analysis of incident disease, we excluded 215 and 91 participants who had prevalent T2D at the time of blood draw from HUNT-Soma5k and HUNT-Soma 7k cohorts, respectively.

### Statistical analysis

Logistic regression analysis was performed to select an obesity predictor with the strongest association with incident T2D to be included in the model. Body mass index (BMI), waist circumference (WC), waist-hip-ratio (WHR), and waist-height-ratio (WHtR) were used individually as proxies for obesity in the logistic regression model with incident T2D as the outcome. Logistic regression analysis between anthropometric measurements and T2D showed that BMI, WC, WHR, and WHtR are comparably associated with T2D as the 95% confidence intervals of the estimated effect sizes are overlapping with WHtR having the slightly larger point estimate (**Supplementary Figure 1**). Therefore, WHtR was chosen to be included in further downstream analyses as the best proxy for obesity.

We used the Kurtze score as physical activity index which was a modified version of the International Physical Activity Questionnaire (IPAQ) to calculate weekly energy expenditure in MET-minutes, focusing on vigorous, moderate, and walking activities. The methods were described in detail elsewhere^(32)^.

To investigate the associations of protein levels with incident T2D, logistic regression analyses were performed for each aptamer. Models included adjustment by sex, age, proxy for obesity, physical activity index, estimated glomerular filtration rate (eGFR), smoking status, and selection criteria (CVD for HUNT-Soma5k and AAA or VTE for HUNT-Soma7k). Model selection was performed by evaluating Nagelkerke’s pseudo-R² statistics across candidate model specifications. The model with protein, sex, age, WHtR, smoking, physical activity, eGFR, and selection criteria had the maximum pseudo-R² in both datasets (**Supplementary Table S1**) and was considered as the final model for further analyses. We chose to analyse HUNT-Soma5k and HUNT-Soma7k separately due to differing selection criteria in the two datasets. We compared multiple model scenarios in which different covariates were added to evaluate how the covariates have associations with the proteins in HUNT-Soma5k (**Supplementary Table S2**) and HUNT-Soma7k (**Supplementary Table S3**). We defined the significant proteins after adjusting for multiple comparisons using Benjamini-Hochberg method with adjusted p-value less than 0.05. Similarly, we investigated the association of protein levels with prevalent T2D using the same covariates in a logistic regression for each protein individually.

We systematically compared proteins which have statistically significant association with incident T2D with three previous studies using different cohorts, such as Atherosclerosis Risk in Communities (ARIC)^(17)^, Cardiovascular Health Study (CHS)^(15)^, and Multiethnic Cohort Study (MEC)^(33)^. These three cohorts will be later referred as replication cohorts (**Supplementary Table S4**). We considered replication in other cohorts when the significant proteins were present in both HUNT-Soma dataset, significant (p-value < 0.05) in at least one of the replication cohorts with consistent direction of effect. A review of the evidence for potentially novel proteins associated with incident T2D was performed via systematic search of the T2D Knowledge Portal^(34)^ and on PubMed with the name of the gene and “diabetes.” A search for genome-wide association study evidence for these genes was done on the GWAS Catalog^(35)^ as of 1 January 2026.

### Protein Risk Score Derivation

Least Absolute Shrinkage and Selection Operator (LASSO), a machine learning method, was implemented to create a protein risk score (ProtRS) for T2D. Protein prediction models were derived with R package *glmnet*^(36)^. Both HUNT-Soma5k and HUNT–Soma7k datasets were split using the function ‘initial_split’ in R package based on T2D incident cases to keep a balanced case/control ratio in training and testing; with 70% of the data used as training data for model fitting and 30% of the data used for model testing. To avoid data leakage, we performed kNN imputation as described previously in training and test sets separately. Important features in each model, then, were identified by the LASSO model with 5-fold cross validation of the training data based on the 1 standard error (SE) l value. The penalty parameter in the LASSO model was adjusted based on the training data to ensure the forced inclusion of established covariates as well as a PGS for T2D, alongside the protein variables. Five LASSO models: (1) only proteins, (2) proteins with covariates, (3) proteins + covariates + T2D PGS, (4) only covariates, and (5) only covariates + T2D PGS were performed. The area under the receiver operator curve (AUC) of the test set was used to assess the quality of discrimination for blood plasma proteome in predicting T2D, using incident T2D data as the outcome. ROC curves were generated with R package ‘*pROC*’^(37)^ and bootstrapped confidence intervals were used. Pairwise DeLong’s test was conducted to compare the AUCs from these models.

All analyses were performed using R version 4.3.3.

### Enrichment Pathway Analysis

Gene set enrichment analysis (GSEA) was used to determine which pathways were significantly enriched in T2D-associated proteins. All proteins were ranked according to their standardized effect size for the T2D association, weighted by the signal-to-noise ratio. Here, signal-to-noise ratio is defined as the mean effect size divided by the standard deviation. Enrichment was assessed using Gene Ontology Biological Process (GO-BP) terms and KEGG pathways. Gene sets were restricted to those containing between 10 and 500 genes. All analyses were conducted in R using the *clusterProfiler*^(38)^ package, and pathways with a Benjamini-Hochberg adjusted p-value below 0.05 were considered as statistically significant.

### Ethical considerations

This present study received ethical approval from the Regional Committee for Medical and Health Research Ethics (REK No. 599896) and project permission from the HUNT Data Access Committee (2024/100799). Informed consent was obtained from all participants.

### Data and Resources Availability

The Trøndelag Health Study (HUNT) may be accessed by application to the HUNT Research Centre at https://www.ntnu.edu/hunt/data. The Trøndelag Health Study (HUNT) has invited persons aged 13 - 100 years to four surveys between 1984 and 2019. Comprehensive data from more than 140,000 persons having participated at least once and biological material from 78,000 persons are collected. The data are stored in HUNT databank and biological material in HUNT biobank. HUNT Research Centre has permission from the Norwegian Data Inspectorate to store and handle these data. The key identification in the data base is the personal identification number given to all Norwegians at birth or immigration, whilst de-identified data are sent to researchers upon approval of a research protocol by the Regional Ethical Committee and HUNT Research Centre. To protect participants’ privacy, HUNT Research Centre aims to limit storage of data outside HUNT databank and cannot deposit data in open repositories. HUNT databank has precise information on all data exported to different projects and can reproduce these on request. There are no restrictions regarding data export given approval of applications to HUNT Research Centre. For more information see: http://www.ntnu.edu/hunt/data.

## Results

### Characteristics of the cohort

Despite being recruited for different outcomes of interest, the main characteristics of HUNT-Soma5k and HUNT-Soma7k are similar with 226 participants overlapping between two datasets. The average age of both datasets was 64 years. There were slightly more men than women in both datasets. In terms of anthropometric measurements, the mean BMI and mean waist circumference value indicated that the participants were overweight (defined as BMI > 25 kg/m^2)^ and had abdominal obesity (waist circumference > 90 cm(39)) (**Table 1**). The proportion of participants reporting having ever smoked exceeded that of never smoking in both datasets.

**Table 1.** Baseline clinical characteristics.

| Baseline characteristics | HUNT-Soma5k<br>5,214 aptamers<br>(n = 3,221) | HUNT-Soma7k<br>7,289 aptamers<br>(n = 2,181) |
| --- | --- | --- |
| Age (years)<sup>a</sup> | 64 ± 10 | 64 ± 13 |
| Sex (% men)<sup>c</sup> | 1954 (61%) | 1203 (55%) |
| BMI (kg/m<sup>2</sup>)<sup>a</sup> | 27.99 ± 4.26 | 28.09 ± 4.56 |
| Waist circumference (cm)<sup>a</sup> | 97.27 ± 11.13 | 97.13 ± 10.97 |
| Hip circumference (cm)<sup>a</sup> | 104.4 ± 6.83 | 103.8 ± 7.45 |
| Physical activity index<sup>b</sup> | 1.90 (1.87) | 2.00 (1.87) |
| Ever smoked<sup>c</sup> | 2141 (66%) | 1417 (65%) |
| CVD selection criteria<sup>c</sup> | 737 (23%) | NA |
| AAA selection criteria<sup>c</sup> | NA | 263 (12%) |
| VTE selection criteria<sup>c</sup> | NA | 867 (40%) |
| Prevalent T2D<sup>c</sup> | 215 (7%) | 91 (4%) |
| Incident T2D<sup>c</sup> | 324 (10%) | 176 (8%) |

*The values are: a=mean (SD); b=median (IQR); c=counts (percentages)*

**Table 2.** Summary statistics of 10 candidate proteins identified from the expanded HUNT-Soma7k panel.

| **Protein Name** | **Gene Symbol** | **Benjamini-Hochberg**<br>**adjusted p-value** | **OR**<br>**(95% CI)** |
| --- | --- | --- | --- |
| G antigen 2B | GAGE2B | 1.84E-04 | 1.54<br>(1.30 – 1.83) |
| Glutathione S-transferase 2 | GSTA2 | 1.35E-03 | 1.45<br>(1.23 – 1.70) |
| Very late Antigen-4 | ITGA4\|ITGB1 | 1.00E-02 | 1.37<br>(1.17 – 1.60) |
| Acyl – CoA - binding domain-<br>containing protein 4 | ACBD4 | 1.17E-02 | 1.30<br>(1.14 – 1.48) |
| Semaphorin-4G | SEMA4G | 1.67E-02 | 1.40<br>(1.17 – 1.70) |
| Chloride intracellular channel protein<br>3 | CLIC3 | 2.99E-02 | 1.32<br>(1.13 – 1.54) |
| Melanoma-associated antigen MUC18 | MCAM | 3.65E-02 | 0.73<br>(0.61 – 0.87) |
| Phosphoserine phosphatase | PSPH | 3.97E-02 | 1.24<br>(1.10 – 1.41) |
| Dr1-associated corepressor | DRAP1 | 4.41E-02 | 1.23<br>(1.09 – 1.39) |
| Pyridoxal phosphate phosphatase 2 | PHOSPHO2 | 4.76E-02 | 1.23<br>(1.09 – 1.39) |

### Proteins associated with T2D

We identified 313 aptamers (308 proteins) from HUNT-Soma5k and 142 aptamers (128 proteins) from HUNT-Soma7k that are significantly (adjusted p-value < 0.05 after FDR correction) associated with incident T2D after adjusting for sex, age, WHtR, eGFR, physical activity, smoking, and selection criteria. The most downregulated proteins were Matrix Remodeling Associated 8 [MXRA8 (OR=0.51, 95% CI 0.44-0.59)] and Insulin Growth Factor Binding Protein 2 [IGFBP2 (OR=0.51, 95% CI 0.41-0.64)] in HUNT-Soma5k and HUNT-Soma7k, respectively (**Figure 2**). Meanwhile, we observed Plexin B2 [PLXNB2 (OR=1.70, 95% CI 1.50-1.94)] in HUNT-Soma5k and Inhibin Subunit Beta C [INHBC (OR=1.96, 95% CI 1.58-2.44)] in HUNT-Soma7k as the most upregulated proteins (**Supplementary Table S5**).

**Figure 1.**
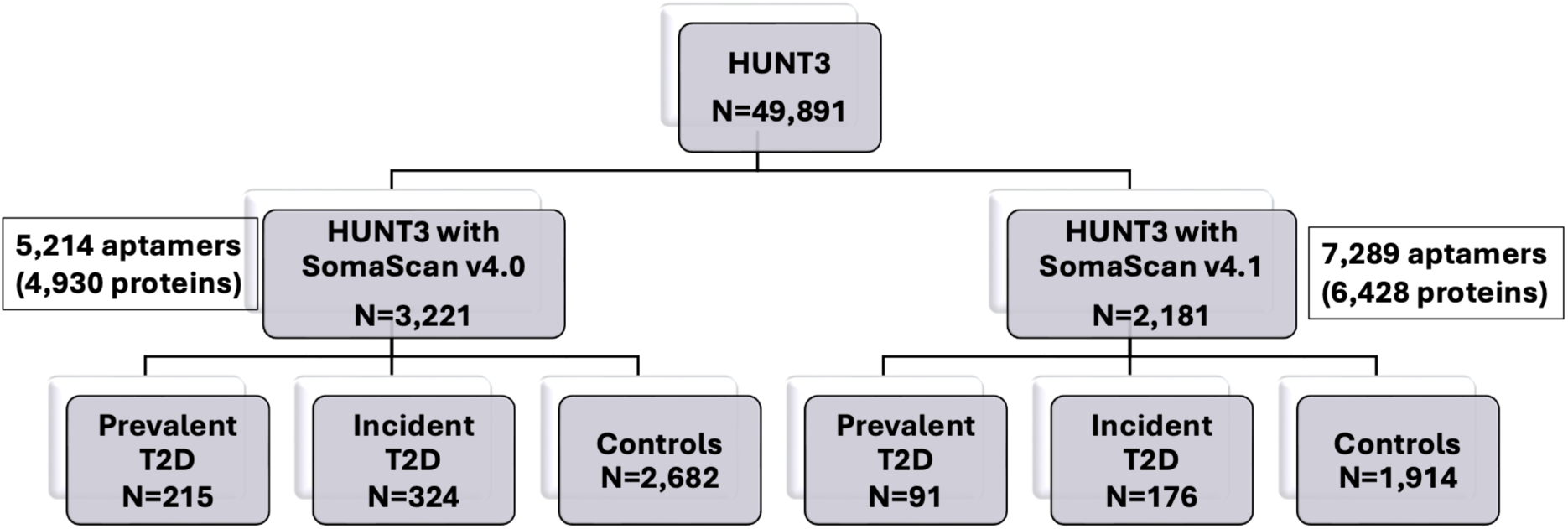
Study design

**Figure 2.**
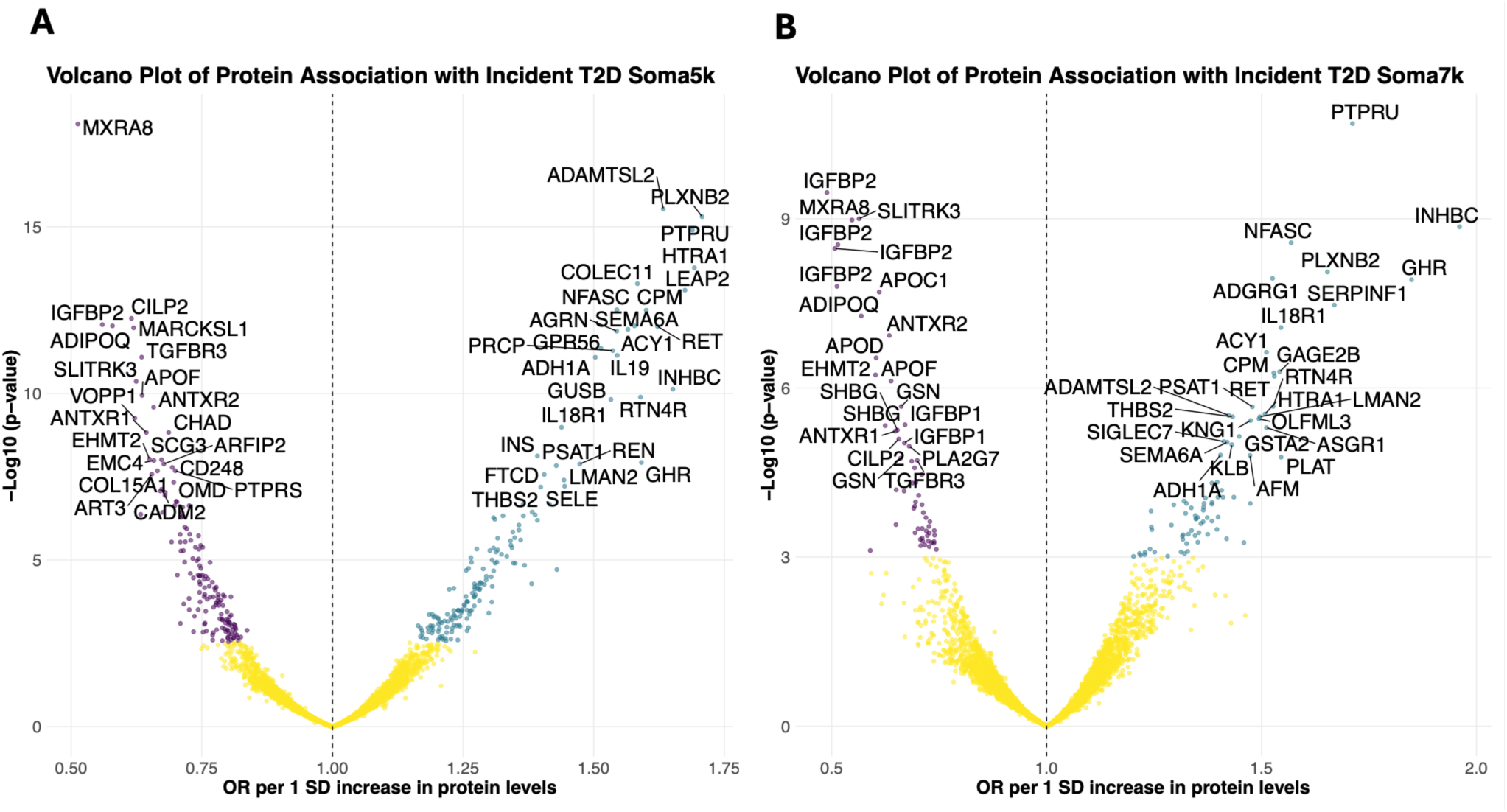
Blood plasma top 50 protein associations for incident T2D in HUNT-Soma5k (**A**) and HUNT-Soma7k (**B**) datasets adjusted for sex, age, WHtR, physical activity, eGFR, smoking, and selection criteria. Volcano plots demonstrating positive (green) and negative (purple) serum associations with T2D incident, points are colored dark where adjusted P-value after FDR<0.05; WHtR=waist-height-ratio, eGFR=estimated glomerulus filtration rate

IGFBP2 is one of the most significantly differentially expressed proteins which have been previously associated with incident T2D^(40)^ and thereby serves as a positive control. The aptamer targeting IGFBP2 in HUNT-Soma5k and HUNT-Soma7k yields similar effect sizes in both cohorts (HUNT-Soma5k OR=0.56, 95% CI 0.48-0.66; HUNT-Soma7k OR=0.51, 95% CI 0.41- 0.64) and replicates with consistent direction of effect in all three replication cohorts (ARIC, CHS, and MEC). Three additional aptamers targeting IGFBP2 in the HUNT-Soma7k are significant with similar effect sizes but were not tested in the replication cohorts (**Supplementary Table S6**).

In addition to analyses in incident T2D, we also tested for proteins associated with prevalent T2D (**Supplementary Table S7, Supplementary Figure 3**), however only incident T2D is considered as the outcome for further downstream analyses. We found 531 (524 proteins) and 335 (306 proteins) aptamers significantly associated with prevalent T2D and among those proteins, 271 (267 proteins) and 123 (112 proteins) aptamers were also significantly associated with incident T2D in HUNT-Soma5k and HUNT-Soma7k, respectively. PLXNB2 was the most upregulated protein associated with both prevalent and incident T2D, while Cartilage Intermediate Layer Protein 2 (CILP2) was the most downregulated in both.

We found 239 and 86 aptamers significantly associated with incident T2D in HUNT-Soma5k and HUNT-Soma7k, respectively, which also replicated in at least one of the replication cohorts (**Supplementary Table S8**). When considering the 10 proteins with the largest positive association with incident T2D in HUNT-Soma5k and HUNT-Soma7K, INHBC, Protein Tyrosine Phosphatase Receptor (PTPRU), and Growth Hormon Receptor (GHR) were also in the top 10 most significant proteins in all replication cohorts. PLXNB2 and Proto-oncogene tyrosin-protein kinase receptor (RET) were in the top 10 most significant proteins in 2 of the replication cohorts. Of the 10 proteins with largest negative association with incident T2D, MXRA8, IGFBP2, Adiponectin (ADIPOQ), and SLIT and NTRK-like family member 3 (SLITRK3) were in both datasets and replicated in all replication cohorts.

We observed a moderate correlation between effect sizes of HUNT-Soma5k and HUNT-Soma7k (Pearson’s correlation of r = 0.62). The regression slope of 0.694 indicates that, on average, effect sizes in HUNT-Soma5k were slightly attenuated compared with HUNT-Soma7k (**Figure 3**), although the overall direction and relative ranking of protein associations were largely preserved. Several proteins, including INHBC and GHR, showed strong consistent positive associations in both datasets, while ADIPOQ and IGFBP2 showed consistent negative associations.

**Figure 3.**
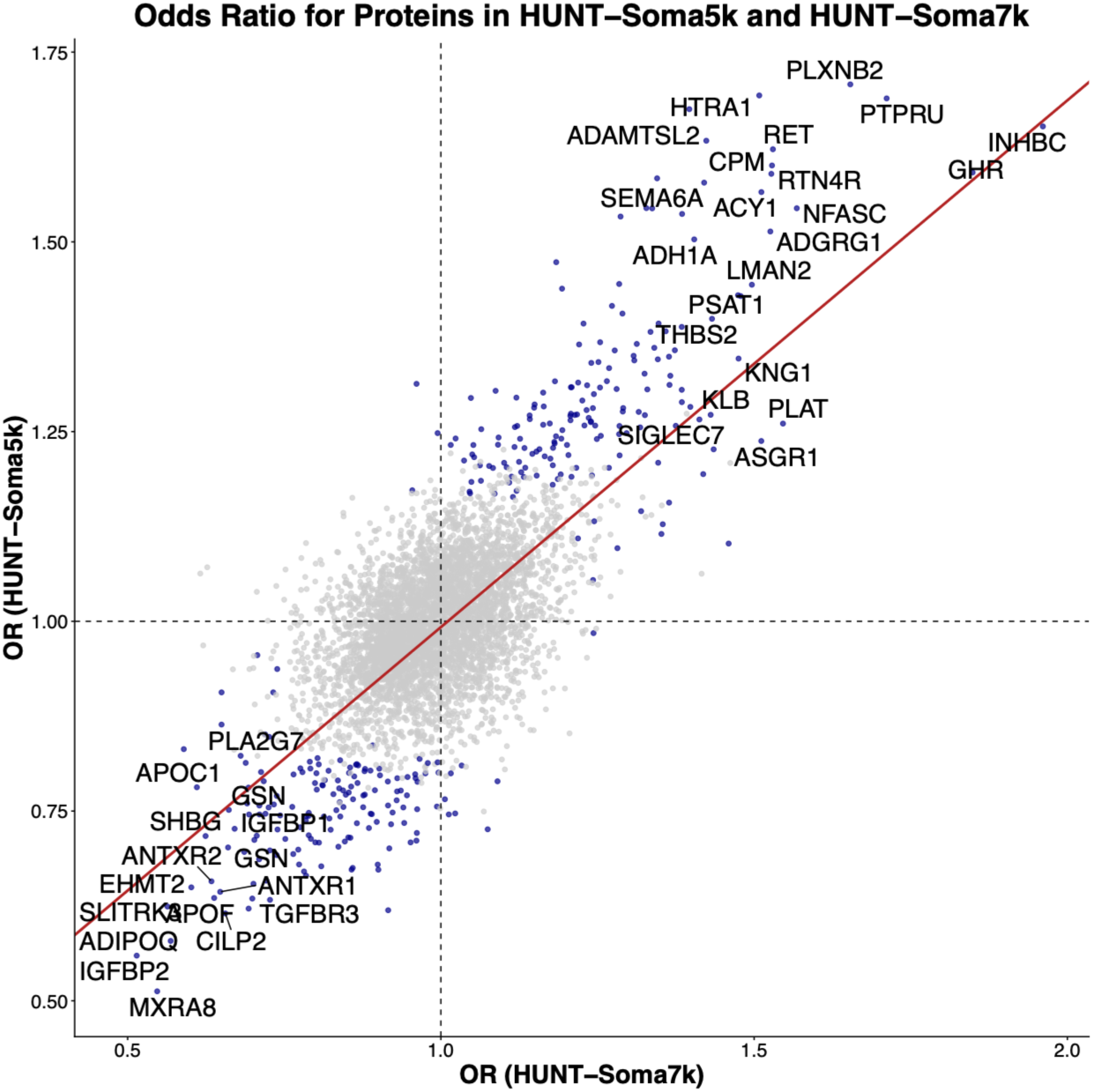
Odds ratio estimates for proteins in HUNT-Soma5k and HUNT-Soma7k. Each point in the scatter plot represents a protein, with its OR estimated independently in each dataset. The fitted regression line, which is shown as the red diagonal line had a slope of 0.694 suggesting moderate concordance in effect sizes across two SomaScan versions.

We identify 38 aptamers (36 proteins) that are statistically significant (FDR < 0.05) and unique to HUNT-Soma7k which used the SomaScan v4.1 assay and are therefore unable to be replicated from previous studies which all used the SomaScan v4.0 assay used in HUNT-Soma5k. Of these, 26 proteins have other aptamers tested in SomaScan-5k and we replicated 11 of these proteins in the replication cohorts. We consider 10 significant proteins for which there is no other evidence in external studies or HUNT-Soma5k as potentially candidate proteins associated with T2D. These proteins are GAGE2B, GSTA2, ITGA4|ITGB1, ACBD4, SEMA4G, CLIC3, MCAM, PSPH, DRAP1, PHOSPHO2 and should be tested in future studies (**Supplementary Table S9**). Evidence from systematic characterization for these 10 proteins is described in the Supplementary Note.

### Proteomic risk score predicts incident T2D

We applied a supervised machine learning method to derive a ProtRS and evaluate its ability to predict incident T2D across two datasets (**Supplementary Figure 3-4**). Models incorporating proteomics showed promising discrimination, with AUCs ranging from 0.79 and 0.84 in both datasets (**Figure 4, Supplementary Table S10**). In contrast, covariates-only and covariates + PGS models showed lower AUCs, consistent with the limited predictive performance of traditional clinical risk factors alone.

**Figure 4.**
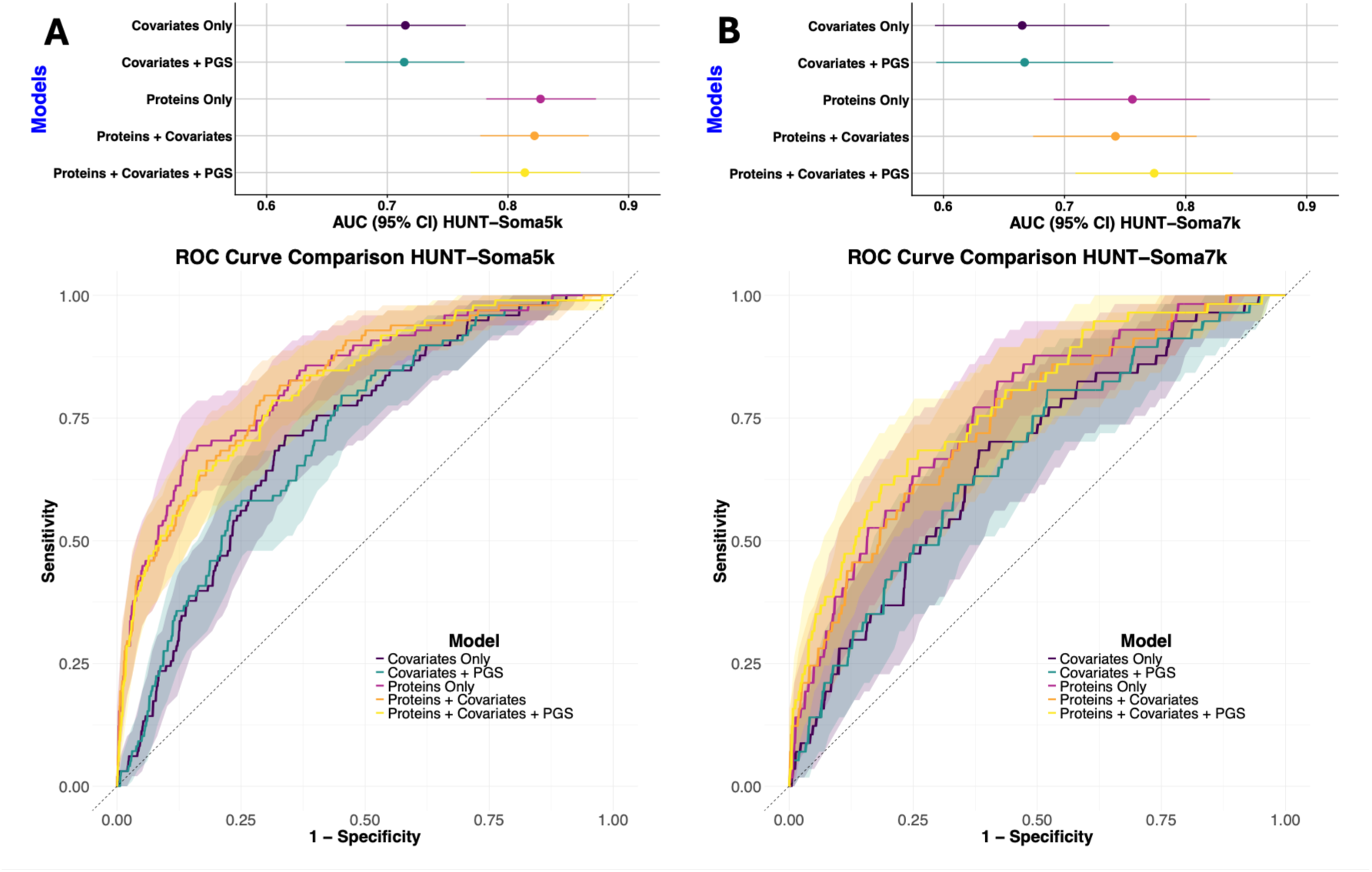
Area under curve (AUC) and ROC curve comparison in HUNT-Soma5k (**A**) and HUNT-Soma7k (**B**) after LASSO indicated that the blood plasma proteome has good discrimination for predicting incident T2D; covariates: sex, age, WHtR, physical activity, eGFR, smoking, and selection criteria; ROC=receiving operating characteristic, WHtR=waist-height-ratio, eGFR=estimated glomerulus filtration rate

To formally compare model performance, we applied DeLong’s test to compare AUCs across models. In the HUNT-Soma5k data, the three models containing proteomics (proteins-only, proteins + covariates, proteins + covariates + PGS) showed no statistically significant differences from one another (**Supplementary Table S11**), indicating that the addition of covariates and/or PGS did not improve discrimination beyond the proteomic signal. However, the AUCs in the three models including proteins compared with only covariates and covariates+PGS models demonstrated statistically significant differences. For instance, the proteins only (p-value = 2.17E-06), proteins+covariates (p-value = 1.56E-09), and proteins+covariates+PGS models (p-value=2.91E-03) showed significantly higher AUCs compared with covariates-only model.

A similar pattern of no statistically significance differences in AUCs between the models including proteins models was observed in the HUNT-Soma7k; and statistically significance differences between only covariates model and the models including proteins: only proteins (p-value = 9.19E-05), proteins+covariates (p-value = 9.93E-08), and proteins+covariates+PGS models (p-value= 1.22E-03). However, the prediction performance in HUNT-Soma7k data set did not improve discrimination when PGS was added to covariates only model compared to models including proteins, except for proteins + covariates + PGS (p-value = 7.31E-06), indicating the importance of proteomics in discriminating performance.

The covariates-only and covariates + PGS models did not differ significantly from one another in both datasets (HUNT-Soma5k p-value = 9.89E-01; HUNT-Soma7k p-value = 3.72E-01). These findings may suggest that the proteins drive the majority of the discriminative ability for predicting incident T2D, with minimal contribution from genomics or covariates.

A previous study^(41)^ using SomaScan platform assay version 3.2 which included 1,095 aptamers, identified 10 proteins associated with incident T2D (**Supplementary Table S12**). Five of those proteins were then included in a LASSO model that did not significantly improve upon a clinical risk score for Diabetes (German Diabetes Risk Score [GDRS]). From those five proteins, four are significant in HUNT-Soma5k and three in HUNT-Soma7k. Only TGFBR3 is included in the present ProtRS for HUNT-Soma5k model (**Supplementary Figures 4B**). However, in the present study, proteomics significantly improved the model when compared to only the clinical risk factors (DeLong’s test p-value = 8.75E-09, **Supplementary Table S5**), many of which are included in the GDRS (e.g. smoking, physical activity). This highlights that the larger set of proteins identified in our study is necessary to improve prediction over clinical risk factors.

### Metabolic processes enriched in genes coding for proteins associated with incident T2D

We identified 41 and 48 significant pathways for Gene Ontology Biological Processes (GO:BP) in HUNT-Soma5k and HUNT-Soma7k, respectively **(Figure 5)**. Organic acid metabolic process showed the most significant association in both HUNT-Soma5k (adjusted p-value=5.33E-07) and HUNT-Soma7k (adjusted p-value=6.30E-06), indicating consistent enrichment of genes associated with this process (**Supplementary Table S13**). Beyond organic metabolic process, our findings also highlighted carboxylic acid metabolic process and oxoacid metabolic process as the top three most statistically significant pathway analysis in HUNT-Soma5k with similar significance (adjusted p-value=2.45E-06), whereas inflammatory response and oxoacid metabolic process are the top three in HUNT-Soma7k (adjusted p-value=6.30E-06; 4.37E-05).

**Figure 5.**
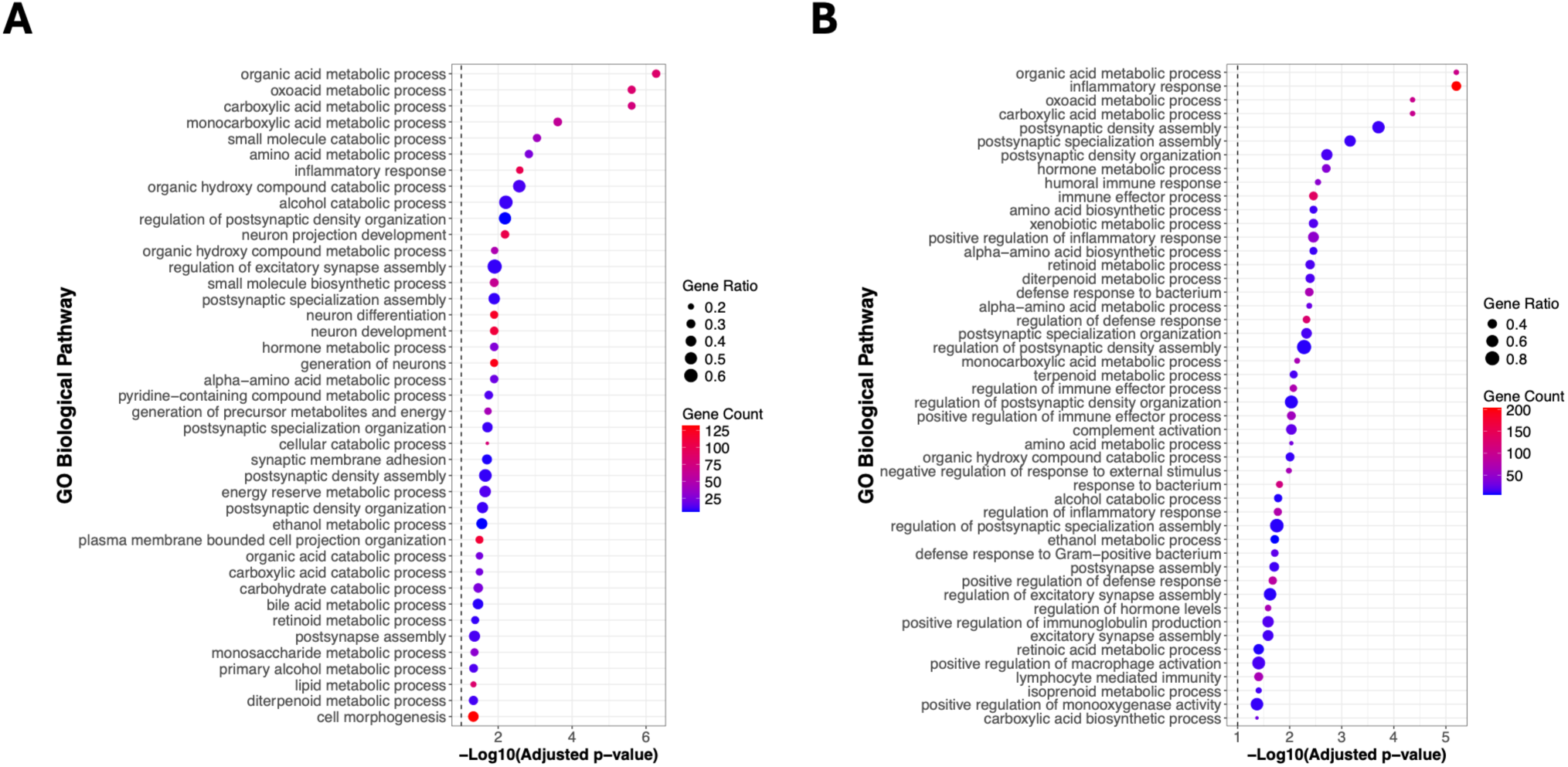
GSEA analysis. Dotplots for GSEA analysis referring to Gene Ontology Biological Process in HUNT-Soma5k (**A**) and HUNT-Soma7k (**B**) showed that organic acid metabolic process as the top biological pathways

In addition to the GO:BP results, we identified 4 and 25 significantly enriched pathways in the KEGG analysis for HUNT-Soma5k and HUNT-Soma7k, respectively. The most statistically significant pathways were pentose and glucoronate interconversions (adjusted p-value = 1.67E-02) in HUNT-Soma5k and metabolism of xenobiotics by cytochrome P450 (adjusted p-value = 2.08E-03) in HUNT-Soma7k, each demonstrating strong enrichment signals relative to other KEGG terms (**Supplementary Table S14, Supplementary Figure 6**). Beyond these top pathways, pancreatic secretion (adjusted p-value=1.67E-02) and IgSF CAM signalling (adjusted p-value=2.03E-02) emerged among the most enriched KEGG pathways in HUNT-Soma5k, whereas retinol metabolism and complement and coagulation were the most prominent pathways in HUNT-Soma7k with similar significance (adjusted p-value=2.08E-03). Taken together, these pathway enrichments suggest a coordinated shift in core metabolic network activity rather than isolated changes in individual proteins. The consistent signal across organic acid, carboxylic acid, and oxoacid metabolism is compatible with early disturbance of mitochondrial substrate flux and intermediary metabolism in the transition toward T2D.

## Discussion

In this study, we identified 83 proteins significantly associated with incident T2D between the two proteomics data sets. In HUNT-Soma5k, PLXNB2 has the strongest positive association with incident T2D. PLXNB2 is a receptor involved in semaphorin signalling and cellular adhesion, and tissue remodelling. The protein is present across metabolic and endocrine tissues, including the thyroid gland, stomach, liver, pancreas, kidney, and adipose tissue^(42)^. A proteomic study in AGES-Reykjavik study has shown a causal link between PLXNB2 and T2D through two-sample MR analyses^(16)^. In contrast, MXRA8 demonstrated the strongest negative association with incident T2D in this dataset. MXRA8 is an extracellular matrix–associated adhesion molecule expressed in several human metabolic tissues, including adipose tissue, liver, and pancreas^(43)^. The protein may play a role in extracellular matrix organization and adipose tissue remodelling, processes that are tightly linked to metabolic homeostasis^(43)^. In the context of incident T2D, the observed downregulation of MXRA8 may reflect impaired adipose tissue remodelling or reduced extracellular matrix turnover.

In HUNT-Soma7k, the INHBC gene which encodes β-C subunit of the inhibin/activin family, forming activin C (Act-C) was observed to have the strongest positive association with incident T2D. This is a hepatokine involved in liver–adipose endocrine signalling^(44)^. A recent bidirectional MR study demonstrated causal relationships between circulating INHBC and multiple cardiometabolic traits, including adiposity, insulin resistance, and systemic inflammation^(45)^. Conversely, IGFBP2 exhibited the strongest negative association with incident T2D in this dataset. IGFBP2 is a circulating insulin-like growth factor–binding protein that modulates IGF bioavailability and metabolic signalling^(46)^. Multiple prospective human cohort studies consistently show that higher circulating IGFBP2 levels are associated with a lower risk of developing T2D, independent of traditional metabolic risk factors^(47, 48)^.

### Candidate proteins identified from HUNT-Soma7k panel associated with incident T2D

We measured abundance for 2,303 aptamers, representing 2,225 proteins, that are unique to the SomaScan v4.1 assay, used in HUNT-Soma7k, compared to SomaScan v4.0. From these, 38 aptamers were statistically significant (Benjamini-Hochberg adjusted p-value < 0.05) and 10 of these measured proteins which were unique to the SomaScan-v4.1 assay (**Supplementary Table S5**). Due to the limited availability of replication data from this assay, we propose these as candidate proteins associated with incident T2D which should be explored in future studies. The genes that encode GSTA2, ACBD4, and PSPH have been associated with T2D in the GWAS Catalog, whereas the gene encoding MCAM and ITGB1 have been associated with HDL and triglycerides levels. However, other proteins such as ITGA4, SEMA4G, CLIC3, DRAP1, and PHOSPHO2 have not been previously linked to T2D^(35)^. The gene that encodes GAGE2B has not been found as the closest gene in the studies on the GWAS Catalog.

ACBD4 was positively associated with incident T2D in our study and the gene encoding the protein, ACBD4, has been previously associated with T2D and BMI-adjusted for hip circumference through genome-wide association studies (GWAS)^(49)^, although no additional evidence was found in the literature describing the mechanism. The Human Genetic Evidence (HuGE) score^(50)^ for the gene provides very strong evidence for association with T2D. The CLIC3 protein was positively associated with incident T2D and the gene encoding CLIC3 has moderate evidence for a range of anthropometric, cardiovascular, glycaemic, hepatic, and lipid traits as estimated by the HuGE score. The 3D chromatin maps suggest CLIC3 as a novel effector gene in the pathogenies of T2D in acinar cells implicated by variant to gene to cell type mapping ^(51)^.

MCAM, also known as CD146, is a known marker for endothelial damage, involved in inflammation and vascular damage. Expression of the CD416 protein has been positively associated with T2D and severity of diabetic nephropathy^(52)^. In our study, MCAM was negatively associated with incident T2D. This may reflect early endothelial dysfunction or reduced vascular resilience which happens in the early stages of diabetes rather than the increased MCAM seen in established diabetes with vascular complications. These novel proteins provide the basis for testing of mechanistic hypotheses and should be replicated in future large-scale proteomics studies of incident diabetes.

### An improved proteomic predictor for T2D

The predictive performance of LASSO-based models incorporating proteomic, clinical, and genetic information for incident T2D indicated strong discrimination potential of the plasma proteome. We observed that proteomic models—whether used alone or combined with covariates and PGS—consistently outperformed models based solely on traditional clinical risk factors. Moreover, the lack of significant differences among the three proteomic models suggests that the proteomic signature itself captures most of the discriminative signal relevant for T2D risk.

Our findings suggest that broader proteomic coverage is essential for improving prediction of incident T2D beyond established clinical risk factors. Earlier work using a smaller aptamer panel^(41)^ reported limited incremental value over the GDRS, whereas our models—built on substantially expanded proteomic profiles—demonstrated clear gains in predictive performance. The partial overlap in significant proteins between studies highlights both shared biology and platform-specific differences, but the improved discrimination observed in this present study indicates that larger, more comprehensive proteomic assays may capture additional biological variation relevant for early T2D development.

### Potential key biological pathways indicated in incident T2D pathogenesis

We identified GO:BP pathways that encompass key enzymatic steps in the handling of amino acids, fatty acids, and TCA-cycle–derived intermediates. Altered abundance of proteins involved in these processes is consistent with prior metabolomic and proteomic studies showing that disruptions in branched-chain amino acid (BCAA) catabolism^(53)^, fatty acid oxidation^(54)^, and ketoacid/TCA-cycle flux^(55)^ precede the onset of insulin resistance and predict incident T2D. Elevated of BCAA-derived ketoacids and other oxoacids have been reported to be linked to hyperinsulinemia and beta cell exhaustion, impaired mitochondrial function and incomplete substrate oxidation^(56)^, while dysregulated carboxylic acid metabolism reflects early defects in lipid handling and energy metabolism^(57)^. These enrichments are consistent with early network-level metabolic dysregulation rather than isolated biomarker effects. In particular, the convergence on organic acid, carboxylic acid, and oxoacid metabolism is compatible with altered mitochondrial substrate handling, perturbed branched-chain amino acid catabolism, incomplete fatty acid oxidation, and overflow of TCA-linked intermediates, all of which have been implicated in the transition from insulin resistance to overt T2D. Framing these proteins as components of an interconnected metabolic response may better capture their biological relevance than considering them individually.

### Strengths and limitations

Our findings should be interpreted in the context of study limitations. First, the relatively limited number of incident T2D cases may have reduced statistical power to detect weaker associations. Second, the HUNT study is predominantly of European ancestry and from a specific geographic region in Norway, which may limit the generalizability of the findings to more diverse populations. Third, although we adjusted for several known risk factors as potential confounders, the possibility of residual confounding from unmeasured or unknown variables cannot be ruled out. Future studies in larger, more diverse populations with comprehensive covariate data are needed to validate and extend these findings. The HUNT3 study’s high participation rates (54%) coupled with low emigration from the catchment area enhance the population representativeness of the findings. However, the proteomics data comes from a case-cohort study of CVD and AAA-VTE, which therefore limits the generalizability. The present study also relied on a single train-test split, which could result in overfitting and reduced generalizability to external datasets. Replication in independent datasets will be crucial to confirm robustness and facilitate translation into clinical contexts.

One of the strengths of this present study is the observation of positive controls (e.g. IGFBP2) which has similar effect sizes for the same aptamer in HUNT-Soma5k and HUNT-Soma7k, across different aptamers for the same protein in HUNT-Soma7k, and for the replication studies. Furthermore, this present study has rigorous quality control by the HUNT databank for the variables used. Additionally, two relatively independent subsets of the HUNT study were used for the analyses, which can be considered a type of internal validation. Lastly, the genetic background of the participants in the HUNT study is relatively homogeneous, minimizing the potential impact of genetic variation on protein expression.

This study identified hundreds of proteins associated with incident T2D in the HUNT study which were consistently replicated in at least one external cohort. We also provide the first evidence for association of SEMA4G, CLIC3, DRAP1, and PHOSPHO2 with T2D. The ProtRS derived from these findings demonstrates strong discrimination between incident T2D and controls, suggesting the potential of a proteome-wide prediction metric for T2D screening.

## Code availability

The code for analyses and visualization is available on Zenodo with DOI: https://doi.org/10.5281/zenodo.21839864.

## Data Availability

The Trondelag Health Study (HUNT) may be accessed by application to the HUNT Research Centre at https://www.ntnu.edu/hunt/data. The Trondelag Health Study (HUNT) has invited persons aged 13 - 100 years to four surveys between 1984 and 2019. Comprehensive data from more than 140,000 persons having participated at least once and biological material from 78,000 persons are collected. The data are stored in HUNT databank and biological material in HUNT biobank. HUNT Research Centre has permission from the Norwegian Data Inspectorate to store and handle these data. The key identification in the data base is the personal identification number given to all Norwegians at birth or immigration, whilst de-identified data are sent to researchers upon approval of a research protocol by the Regional Ethical Committee and HUNT Research Centre. To protect participants privacy, HUNT Research Centre aims to limit storage of data outside HUNT databank and cannot deposit data in open repositories. HUNT databank has precise information on all data exported to different projects and are able to reproduce these on request. There are no restrictions regarding data export given approval of applications to HUNT Research Centre. For more information see: http://www.ntnu.edu/hunt/data.

https://doi.org/10.5281/zenodo.21839864.

## Acknowledgment

We want to thank clinicians and other employees at Nord-Trøndelag Hospital Trust for their support and for contributing to data collection in this research project. The Trøndelag Health Study (HUNT) is a collaboration between HUNT Research Centre (Faculty of Medicine and Health Sciences, Norwegian University of Science and Technology NTNU), Trøndelag County Council, Central Norway Regional Health Authority, and the Norwegian Institute of Public Health. The genotyping in HUNT was financed by the National Institutes of Health; University of Michigan; the Research Council of Norway; the Liaison Committee for Education, Research and Innovation in Central Norway; and the Joint Research Committee between St Olavs hospital and the Faculty of Medicine and Health Sciences, NTNU.

Members of the HUNT All-In Research Team (in alphabetical order by surname): Bjørn Olav Åsvold, Ben Brumpton, Maiken Elvestad Gabrielsen, Kristian Hveem, Ida Surakka, Laurent Thomas, Brooke N. Wolford, and Wei Zhou.

## Author Contributions

FK performed analyses. BNW conceived of and funded the study. BOA, GG, EA, and ISF provided analytical expertise and data interpretation. FK and BNW wrote the manuscript. All authors provided critical feedback of the manuscript and approved of its submission.

Dr. Brooke N. Wolford is the guarantor of this work and, as such, had full access to all the data in the study and takes responsibility for the integrity of the data and the accuracy of the data analysis.

## Ethics Statement

The genotyping in Trøndelag Health Study and work presented here was approved by the Regional Committee for Ethics in Medical Research, Central Norway (2014/144, 2018/1622, and 2018/411492). All participants signed informed consent for participation and the use of data in research.

During the course of preparing this work, the author(s) used Microsoft Copilot for the purpose of rewriting sentences for clarity with academic standards and suggest step-by-step fixes for broken code. Following the use of this tool/service, the author(s) formally reviewed the content for its accuracy and edited it as necessary. The author(s) take full responsibility for all the content of this publication.

